# COVID-19: Rapid Antigen detection for SARS-CoV-2 by lateral flow assay: a national systematic evaluation for mass-testing

**DOI:** 10.1101/2021.01.13.21249563

**Authors:** Tim Peto, UK COVID-19 Lateral Flow Oversight Team

**Author notes:** Corresponding Author: Prof Tim Peto.

## Abstract

Lateral flow device (LFD) viral antigen immunoassays have been developed around the world as diagnostic tests for SARS-CoV-2 infection. They have been proposed to deliver an infrastructure-light, cost-economical solution giving results within half an hour. Here we report on standardised laboratory evaluations of LFDs, and for those that met the published criteria, field testing in the Falcon-C19 research study and UK pilots (UK COVID-19 testing centres, hospital, schools, armed forces). 4/64 LFDs so far have desirable performance characteristics (Orient Gene, Deepblue, Abbott and *Innova SARS-CoV-2 Antigen Rapid Qualitative Test*). All these LFDs have a viral antigen detection of >90% at 100,000 RNA copies/ml. 8951 Innova LFD tests were performed with a kit failure rate of 5.6% (502/8951, 95% CI: 5.1-6.1), false positive rate of 0.32% (22/6954, 95% CI: 0.20-0.48). Viral antigen detection/sensitivity across the sampling cohort when performed by laboratory scientists (156/198, 95% CI 72.4-84.3) was 78.8%. Our results suggest LFDs have promising performance characteristics for mass population testing and can be used to identify infectious positive individuals. The Innova LFD shows good viral antigen detection/sensitivity with excellent specificity, although kit failure rates and the impact of training are potential issues. These results support the expanded evaluation of LFDs, and assessment of greater access to testing on COVID-19 transmission.

**Funding:** Department of Health and Social Care. University of Oxford. Public Health England Porton Down, Manchester University NHS Foundation Trust, National Institute of Health Research.

## Introduction

National governments and international organisations including the World Health Organisation (WHO) and European Commission have highlighted the importance of individual testing, mass population testing and subsequent contact tracing to halt the chain of transmission of SARS-CoV-2, the virus responsible for COVID-19.^1,2,3^ The current diagnostic test involves reverse-transcription polymerase chain reaction (RT-PCR) testing of nose/throat swabs in specialised laboratories. Such capacity in the UK is currently estimated at ∼500,000 tests/day^4–7^ and this is used with contact tracing procedures and mobile applications to identify close symptomatic contacts of infected symptomatic individuals.^8–10^ However, there are significant challenges in creating testing capacity to identify those with asymptomatic infections or to test contacts of individuals with COVID-19. To date, turnaround time for RT-PCR has been typically slow (>24 hours).

To better understand and control SARS-CoV-2 transmission, there is an urgent need for large-scale, accurate, affordable and rapid diagnostic testing assays, with the ability to detect infectious individuals. Lateral flow device (LFD) immunoassays can be designed to test for different protein targets and are routinely used in healthcare settings principally as a result of their affordability, ease of use, short turnaround time, and high-test accuracy. In brief, a sample is placed on a conjugation pad where the analyte (or antigen) of interest is bound by conjugated antibodies. The analyte-antibody mix subsequently migrates along a membrane by capillary flow across both ‘test’ and ‘control’ strips. These strips are coated with antibodies detecting the analyte of interest and a positive test is confirmed by the appearance of coloured control and test lines.^11^

Newly developed SARS-CoV-2 antigen LFDs identify the presence of specific viral proteins, using conjugated antibodies to bind spike, envelope, membrane or nucleocapsid proteins. In contrast to the IgM/IgG “antibody tests”, these antigen tests directly identify viral proteins, and are not reliant on the host’s immune response. In contrast to RT-PCR, results for LFDs are observed in 10-30 minutes depending on the device, providing a window for early interventions to halt the chain of transmission earlier in the disease course when individuals are most infectious.^12^

To date, many manufacturers have developed first-generation rapid SARS-CoV-2 antigen-detecting LFDs. However, many of these tests have not been independently validated. There is evidence of variable performance when assessing test sensitivity and specificity, although several candidates looked promising on the basis of early data.^13–15^ An independent national evaluation of these devices is important to facilitate population-level or mass testing initiatives globally.

Here, we report the diagnostic performance of first-generation SARS-CoV-2 antigen-detecting LFD for rapid point-of-care (POC) testing in work that was commissioned by the UK’s Department of Health and Social Care (DHSC) from PHE Porton Down and the University of Oxford.

## Results

### Phase 1

A total of 132 suppliers of SARS-CoV-2 antigen detection LFDs were identified and referred to the DHSC for initial Phase 1 review. Among these, at the time of publication, 64 were selected by the DHSC for further evaluation by the UK lateral flow oversight group.

### Phase 2

As part of Phase 2 evaluations, 9,692 LFD tests were performed at PHE Porton Down across the 64 candidate devices as of the 3^rd^ December 2020. 5 LFDs had a kit failure rate above the pre-specified threshold for exclusion (>10%), 17 kits had a false-positive rate below the pre-defined specificity threshold (<97%) and 28 kits a false-negative rate below the LOD threshold (<60% at 10^2^ pfu/m). In total, across all three criteria, nineteen kits performed at a level in accordance with the UK Lateral Flow Oversight Group’s *a priori* “prioritisation criteria”. All nineteen kits also passed cross-reactivity analyses against seasonal human coronaviruses.

### Phase 3

To date, eight LFDs have passed Phase 3a evaluation, namely: *Innova SARS-CoV-2 Antigen Rapid Qualitative Test* (Innova), *Zhejiang Orient Gene Biotech Co. Coronavirus Ag Rapid Test Cassette* (Swab) (Orient Gene), *Anhui Deepblue Medical Technology COVID-19* (*Sars-CoV-2*) *Antigen Test kit* (*Colloidal Gold*) (Deepblue), *Fortress Diagnostics Coronavirus Ag Rapid Test* (*Fortress*), *Roche SD Biosensor Standard Q COVID-19 Ag Test* (*SD Bio swab*), *Surescreen Diagnostics SARS-CoV-2 Antigen Rapid Test Cassette* (*Nasopharyngeal swab* (*Surescreen*) *and Abbott Panbio COVID-19 Ag Rapid Test Device* (*Abbott*) (Supplementary Table 1). Three LFDs did not pass 3a evaluation and the remaining LFDs are currently undergoing evaluation. Four LFDs (Deepblue, Innova, Orientgene, Abbott) have passed Phase 3b evaluation (Table 1, Supp Figure 1), one LFD did not pass and the remainder have not been evaluated.

**Table 1.** Results of the Phase 3b evaluations showing viral antigen detection/sensitivity of four LFD tests using dry-swab samples from community sampling. Tests were performed by laboratory scientists. Ct – cycle threshold on RT-PCR.

| Viral Load | Average Ct | Innova Number tested/number positive (%) | Abbott Number tested/number positive (%) | Orient Gene Number tested/number positive (%) | Deepblue Number tested/number positive (%) |
| --- | --- | --- | --- | --- | --- |
| >10million | <18 | 5/5 (100) | 1/1 (100) | - | 3/3 (100) |
| 1-10 million | 18-21.5 | 23/23 (100) | 12/13 (92) | 17/17 (100) | 19/19 (100) |
| 0.1-1 million | 21.5-25 | 52/54 (96) | 19/21 (91) | 18/18 (100) | 43/44 (98) |
| 10,000-100,000 | 25-28 | 37/42 (88) | 13/13 (100) | 18/19 (95) | 38/38 (100) |
| 1,000-10,000 | 28-31 | 25/33 (76) | 17/19 (90) | 14/18 (78) | 18/29 (62) |
| 100-1,000 | 31-34.5 | 11/33 (33) | 10/26 (39) | 11/19 (58) | 8/36 (22) |
| <100 | >34.5 | 2/7 (29) | 1/6 (17) | 0/4 (0) | 0/8 (0) |
| Overall | na | 155/197 (79) | 73/99 (74) | 78/95 (82) | 129/177 (73) |

### Extended Innova LFD evaluation (Phases 2-4)

The limit of detection of the Innova LFD (Table 2) was determined as part of Phase 2 evaluations for the Innova test. This analysis consisted of saliva spiked with SARS-CoV-2 with stock of SARS-CoV-2 with a standardised PFU. Under these ideal concentrations, at an estimated PFU of 390/mL, which corresponds to a Ct of ∼25, the LFD identified all samples.

**Table 2.**
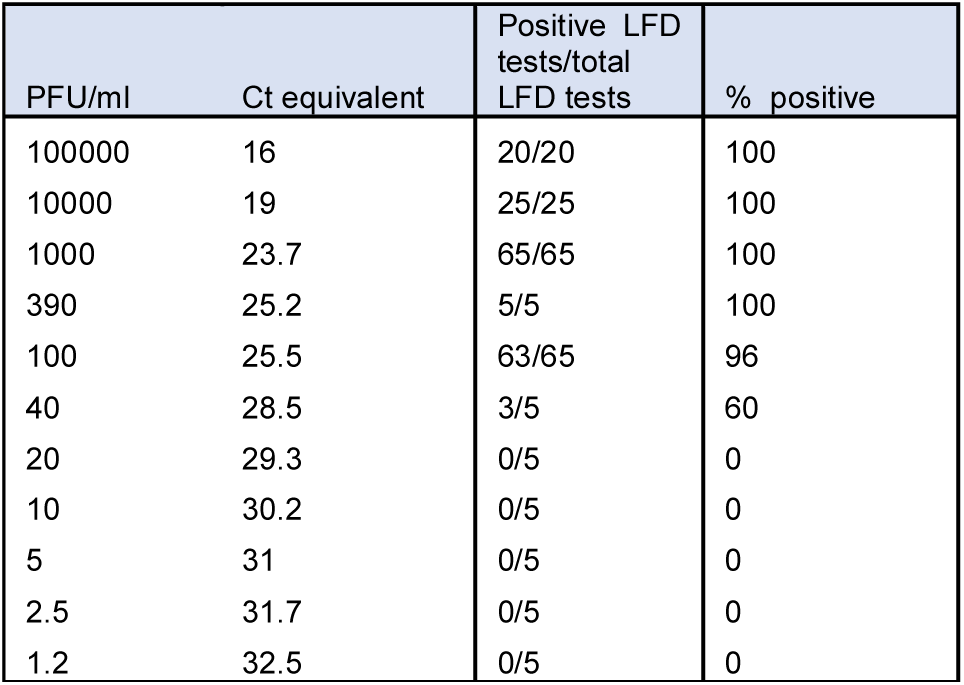
Limit of sensitivity for SARS-CoV-2 detection by the Innova LFD for antigen detection using saliva sample spiked with SARS-CoV-2. Ct - cycle threshold. PFU - plaque forming units.

Our phase 4 evaluation focused on field testing of the Innova LFD, for which we had a sufficient supply of kits available for wider testing at the time. Device specificity was determined through an analysis of 6954 tests from evaluation phases 2-4. The percentage of false-positives ranged from 0.00-0.49%, with an overall specificity of 99.68%. The false-positive rate was centre-dependent (p=0.014, Fisher’s exact test). These evaluations noted that where there were challenges in interpreting the results when the test result was “weak” (i.e. the test line was very faint) (Table 3).

**Table 3.**
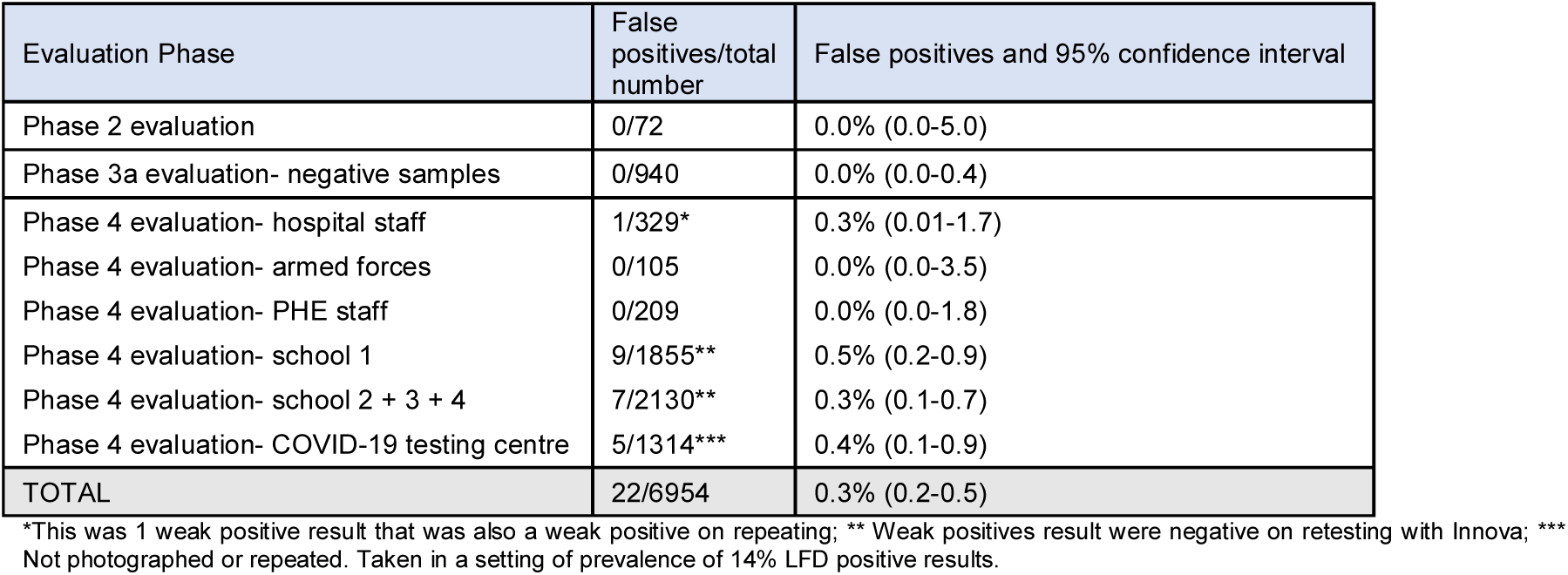
Number of false positives in negative samples in each evaluation stage for the Innova LFD. 95% confidence intervals presented in each case.

| Evaluation Phase | False positives/total number | False positives and 95% confidence interval |
| --- | --- | --- |
| Phase 2 evaluation | 0/72 | 0.0% (0.0-5.0) |
| Phase 3a evaluation- negative samples | 0/940 | 0.0% (0.0-0.4) |
| Phase 4 evaluation- hospital staff | 1/329* | 0.3% (0.01-1.7) |
| Phase 4 evaluation- armed forces | 0/105 | 0.0% (0.0-3.5) |
| Phase 4 evaluation- PHE staff | 0/209 | 0.0% (0.0-1.8) |
| Phase 4 evaluation- school 1 | 9/1855** | 0.5% (0.2-0.9) |
| Phase 4 evaluation- school 2 + 3 + 4 | 7/2130** | 0.3% (0.1-0.7) |
| Phase 4 evaluation- COVID-19 testing centre | 5/1314*** | 0.4% (0.1-0.9) |
| TOTAL | 22/6954 | 0.3% (0.2-0.5) |

\*This was 1 weak positive result that was also a weak positive on repeating; \*\* Weak positives result were negative on retesting with Innova; \*\*\* Not photographed or repeated. Taken in a setting of prevalence of 14% LFD positive results.

Across Phase 2-4 evaluation stages, 8,951 Innova LFD tests were performed, including a diverse cohort of populations as part of Phase 3b and Phase 4 testing, namely out-patient SARS-CoV-2 cases, healthcare staff, armed forces personnel and secondary school children. The overall kit failure rate for the Innova LFD was 5.6% (502/8951, 95% CI: 5.1-6.1) (Table 4). The most common reason for kit failure was poor transfer of the liquid within the device from the reservoir onto the test strip.

**Table 4.** Evaluations of the Innova LFD across Phases 2-4. The table demonstrates the kit failure rate.

| Innova LFD evaluation phase | LFD failures (%) |
| --- | --- |
| Phase 2 negatives | 0/72 (0.0%) |
| Phase 2 positive dilution series | 0/60 (0.0%) |
| Phase 2 positive extended dilution series | 0/155 (0.0%) |
| Phase 2 Swab comparison | 0/187 (0.0%) |
| Phase 3a positives | 13/191 (6.8%) |
| Phase 3a negatives | 50/990 (5.1%) |
| Phase 3b FALCON (Dry swabs- field) | 27/267 (10.1%) |
| Phase 3b FALCON (Dry swabs- lab) | 9/212 (4.2%) |
| Phase 3b FALCON (VTM swabs) | 9/157 (5.7%) |
| Phase 4 hospital staff | 17/358 (4.7%) |
| Phase 4 armed forces | 6/157 (3.8%) |
| Phase 4 PHE staff | 19/212 (8.9%) |
| Phase 4 school 1 | 311/1855 (16.8%) |
| Phase 4 school 2 + 3 + 4 | 14/2132 (0.7%) |
| Phase 4 COVID-19 testing centre | 27/1946 (1.4%) |
|  | 502/8951 (5.6%) |

Viral antigen detection/sensitivity in individuals with confirmed SARS-CoV-2 infection using the Innova LFD was assessed in the Phase 3b evaluation as part of the FALCON-C19 research study. Optimal viral antigen detection/sensitivity when performed by laboratory scientists, was 78.8% (95% CI 72.4-84.3%; 156/198 cases where a paired PCR was performed; see below for differing performance by test operator category). Subgroup analyses showed there were no discernible differences in viral antigen detection/sensitivity in those without symptoms vs. symptomatic individuals (27/41 [65.9%] vs. 95/344 [72.4%], p=0.38). We did not find any evidence of associations between LFD positivity and symptoms or past medical history, with the exception of presence of headache (Supplementary Table 2).

The association between Innova LFD viral antigen detection/sensitivity and estimated viral load/Ct value was explored using the paired RT-PCR VTM swab sample taken at the same time as the swab used for LFD. There was a strong association between viral load detection (RNA copies/mL) determined through RT-PCR and viral antigen detection by LFD (Figure 1). Confirming earlier analyses, sensitivity of LFDs is highest in samples with higher viral loads.^18 19^

**Figure 1.**
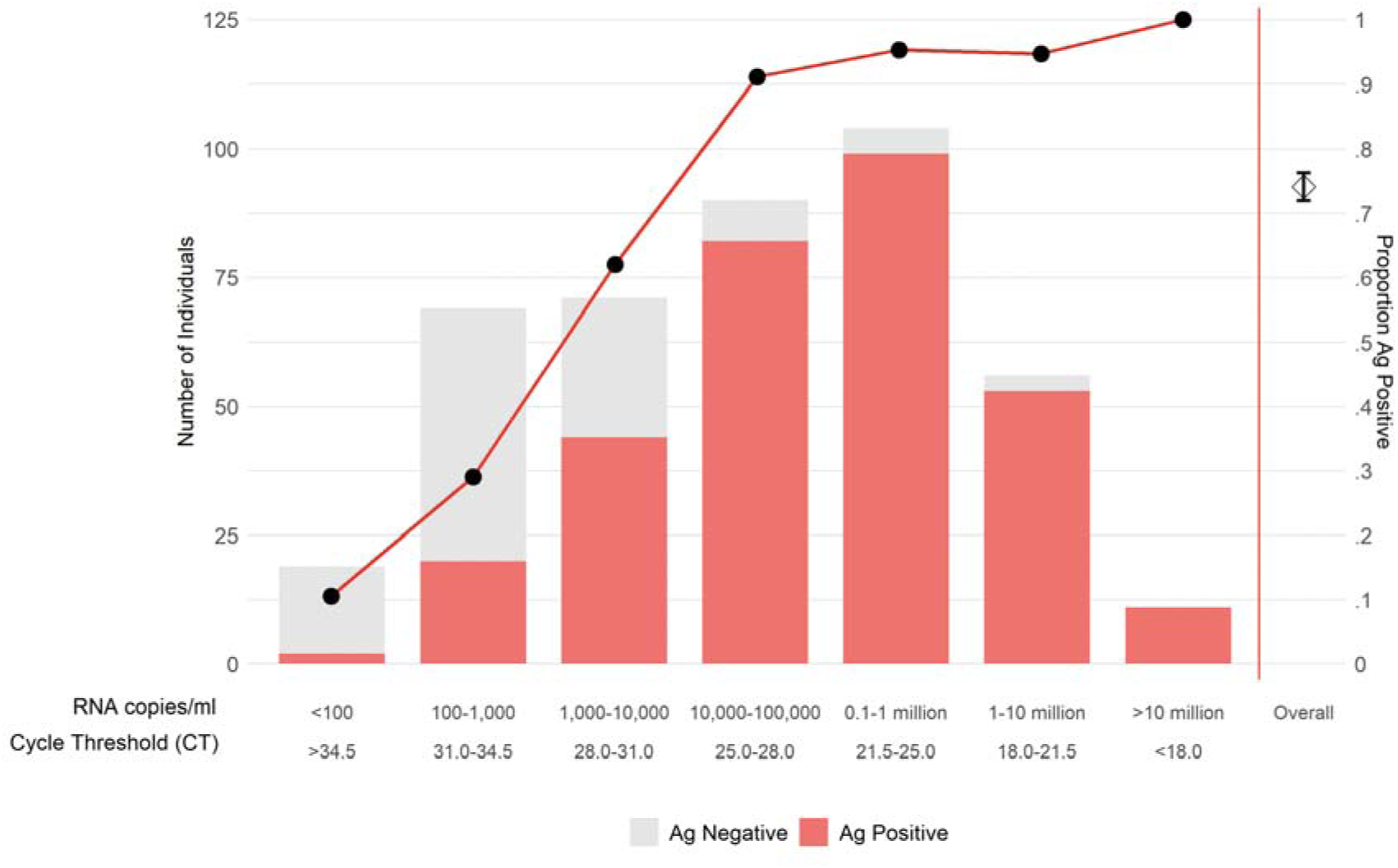
Association between viral antigen detection/sensitivity and viral load (RNA copies/mL and Ct) in Phase 3b Falcon-C19 study evaluation for dry swabs when performed by trained laboratory scientists and trained healthcare workers. Diamond shows point estimate, with 95% confidence intervals, pooling data from all other categories.

**Figure 2.**
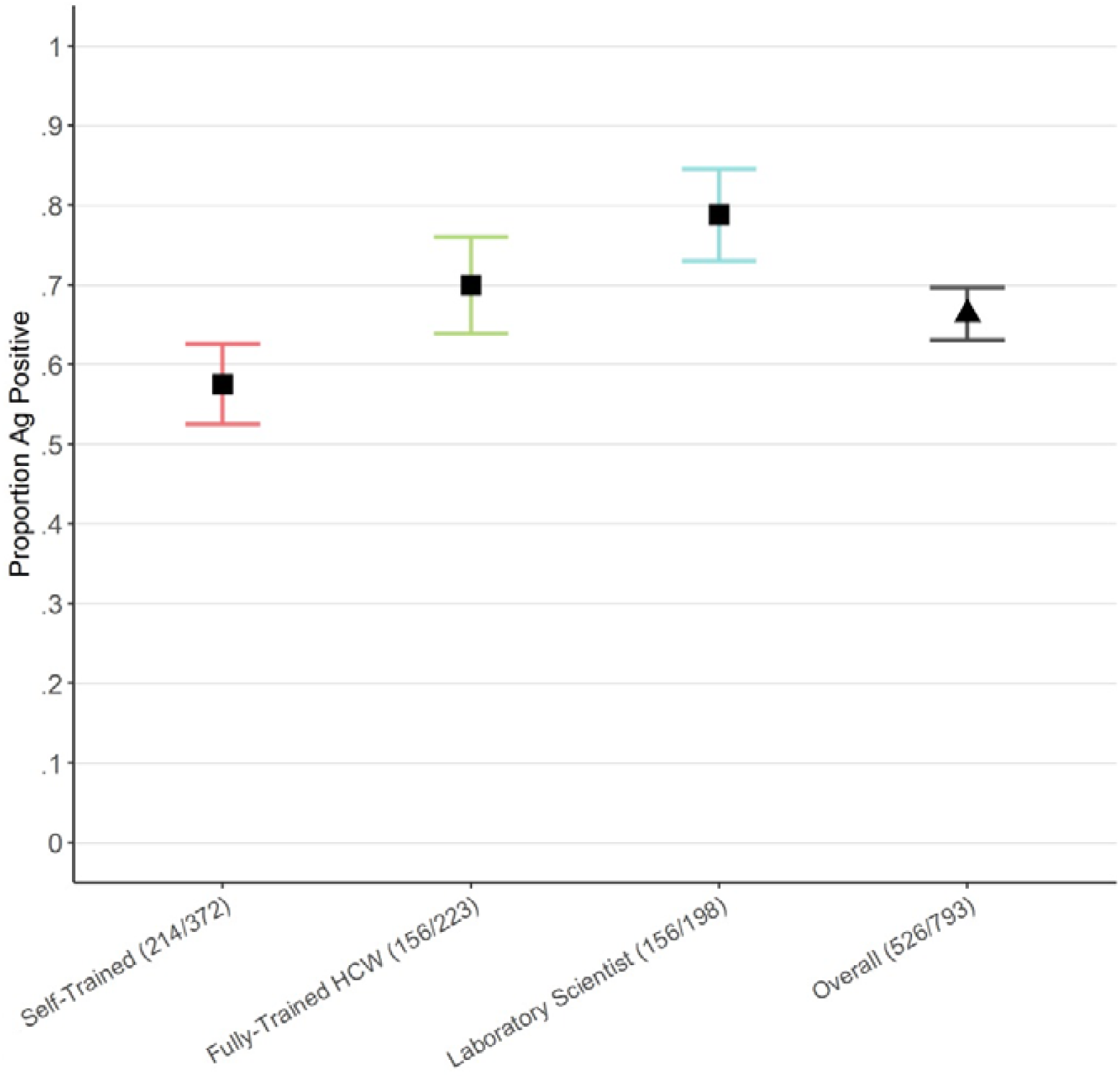
Effect of training and operator on the viral detection/sensitivity of the Innova LFD in COVID-19 PCR-positive patients.

Within the 3b FALCON-C19 study, LFDs were also assessed by sampling 150uL of viral transport medium (VTM) solution instead of using dry swabs; this was associated with poorer performance rate (Supp Figure 2). The use of dry swabs forms the basis of the manufacturer’s instructions for use. This was likely due to a dilution factor involved in placing the swab first into VTM and then analysing the VTM sample, and highlights potential issues in generating direct comparisons between LFDs and VTM samples (Supp Figure 2).

### LFD test performance by operator

As part of Phase 3b-4 evaluations, work was performed to report on the effect of the operator on viral antigen detection/sensitivity in RT-PCR-positive cases using the Innova LFD. Tests were classified according to whether they were performed by a laboratory scientist, a fully trained research health care worker or by a self-trained lay individual working at a regional NHS Test and Trace centre. Performance was optimal when the LFD was used by laboratory scientists (156/198 LFDs positive [78.8%, 95% CI: 72.4-84.3%]) relative to trained healthcare-workers (156/223 LFDs positive [70.0%, 95% CI: 63.5-75.9%]) and self-trained members of the public given a protocol (214/372 LFDs positive [57.5%, 95% CI: 52.3-62.6%]; p<0.0001).

## Discussion

We report on our national evaluation of SARS-CoV-2 viral antigen-detecting LFDs, focussing on the *Innova SARS-CoV-2 Antigen Rapid Qualitative Test*, which has a viral antigen detection (sensitivity) of 78.8% when performed by laboratory scientists and a specificity of 99.7%, using RT-PCR as ‘gold standard’ for positive and negative status. Test performance to detect SARS-CoV-2-positive samples was improved at lower Ct values/higher viral loads, and were >90% at Ct values <25 (equating to ∼390 pfu/mL, 100,000 RNA copies/ml) (Supplementary Table 3). There is an expanding body of evidence that suggests viral load/antigen is important as individuals with the highest viral loads are the most infectious, ^20^ and the presence/absence of viral antigens determined by LFDs is more strongly associated with a viral culture than RT-PCR positivity.^21^ In our evaluation, test performance was largely maintained across different settings and cohorts; however, performance was partly operator-dependent and kit failures are not infrequent.

Our experience is that many LFDs entering our national evaluation program do not perform at a level required for mass population deployment and this reflects the literature. To date, an increasing number of evaluations of SARS-CoV-2 antigen-detecting LFD have been published with variable results. A number of LFDs show good^24 25 13 19 26 27^ or acceptable sensitivity and specificity^28 29^, however, many studies have identified tests with poor sensitivities or specificities.^30 15^

A challenge for most countries during the SARS-CoV-2 pandemic has been the expansion of capacity for diagnostic testing to support the identification of symptomatic and asymptomatic cases. This would aid in offering testing to “contacts” of COVID-19 and enable targeted testing to better safeguard vulnerable populations e.g. care home residents. Reliance on RT-PCR involves significant infrastructural and specialist human resources to implement at increasing scale. Both the World Health Organisation and European commission have issued guidance supporting wider implementation of antigen-targeting LFDs, and in November, Slovakia became the first country in the world to implement entire population testing using LFDs. ^1,3,31^ The UK has similar aspirations to pursue a strategy of mass testing and has implemented a city wide mass testing in Liverpool using the Innova LFD in this study.^32^

It is important to note that there are some potential issues with considering RT-PCR as the gold standard test for COVID-19. Many individuals have persisting viral RNA fragments that can linger for weeks-months without any evidence of active viral replication; in this instance a PCR-positive is likely to overcall the “infectious” status of an individual ^33^ Indeed, when compared to the ability to perform viral culture, data suggest that RT-PCR tends to overestimate the presence of replicating or infectious virions.^34^

In field testing, performance of the Innova LFD was dependent on the test operator. Individuals who had read a protocol immediately prior to self-sampling did not perform as well as individuals with hands-on training, or clinical laboratory personnel who had performed several hundred LFD tests. Like other operator-dependent procedures, further work is required to determine the duration and content of “training” to derive optimal test performance. We also assume that the use of LFDs to successfully identify individuals with higher viral loads and enabling an earlier diagnosis will be of benefit in interrupting transmission, however, this remains to be proven.

SARS-CoV-2 control will benefit from a variety of testing strategies. This might include those optimised for determining past infection/exposure (e.g. serology), those that are of benefit in determining current/recent infection (e.g. RT-PCR), or those identifying potential infectivity. A combination of approaches incorporating the strengths of each of these tests can be effectively used for individuals and for population-level management of the pandemic. Approaches to testing will remain relevant even when effective vaccines become available as it may take several months for an appreciable effect on transmission to be fully realised.^35^

In conclusion, we completed late stage evaluations of seven LFDs. We report sensitivities of 70-80% and specificities ≥99.7% for each LFD evaluated in phase 3b, which involved testing by laboratory personnel or trained healthcare professionals. To identify patients with higher viral loads (Ct<25), each LFD had >90% sensitivity. Sensitivity was lower in phase 4 evaluations, while specificity was maintained. The simplicity of LFDs, without a requirement for specialist training or equipment, mean that they are an attractive option for mass testing. Future research should focus on post-implementation evaluation of diagnostic accuracy, including the potential benefit of regular serial sampling to improve accuracy and reduce transmission.

## Online Methods

A phased evaluation of available SARS-CoV-2 antigen LFDs was undertaken.

### Department of Health and Social Care evaluation (Phase 1 evaluation)

The DHSC identified manufacturers supplying SARS-CoV-2 antigen LFDs that could enable mass testing at a population level. A desktop review was performed to ensure there were appropriate instructions for use and to assess manufacturers’ claimed performance and manufacturing capabilities.^16^

### Pre-clinical evaluation (Phase 2 evaluation)

Pre-clinical evaluation of candidate LFDs was performed by trained laboratory scientists at Public Health England (PHE) Porton Down. LFDs were evaluated against SARS-CoV-2 spiked positive controls and known negative controls, consisting of saliva collected from healthy adult staff volunteers.

Pre-defined and publically available “prioritisation” criteria to pass on to the next evaluation phase had to be met for LFDs, consisting of (i) a kit failure rate of <10%; (ii) an analytical specificity of ≥97%, and (iii) an analytical LOD of ≥9 of 15 (60%) at 10^2^ pfu/mL, corresponding to a RT-PCR cycle threshold (Ct) of approximately 25 (∼100,000 RNA copies/ml); and (iv) lack of cross-reactivity with seasonal coronaviruses to further test analytical specificity.

### Retrospective secondary care evaluation (Phase 3a evaluation)

Evaluation using patient samples retrospectively was started in August 2020 at PHE Porton Down. Samples were obtained from a secondary healthcare setting (Oxford University Hospitals NHS Foundation Trust).

- 1,000 SARS-CoV-2 negative samples: fresh samples held refrigerated were supplied the day after they were tested negative by RT-PCR by the laboratory service at the John Radcliffe Hospital, Oxford, UK.
- 200 SARS-CoV-2 positive samples: swabs collected in VTM from patients admitted to hospital during the first wave of the UK pandemic (March-June 2020).^17^ These were diluted 1:4 SARS-CoV-2 RT-PCR negative saliva, aliquoted and frozen at −20°C for later use. For each positive sample, in addition to the original diagnostic RT-PCR Ct value, a confirmatory RT-PCR was performed at PHE Porton Down on the diluted sample to determine the new Ct value.

### Community research evaluation (Phase 3b evaluation)

We undertook a field evaluation using samples from volunteers in the community in collaboration with the National Institute for Health Research (NIHR) funded CONDOR Platform “COVID-19 National Diagnostic Research and Evaluation Platform”. This was performed within the FALCON-C19 study (Facilitating Accelerated Clinical validation Of Novel diagnostics for COVID-19, 20/WA/0169, IRAS 284229), between 17^th^ September and 23^rd^ October 2020. This involved the recruitment and re-testing of consenting adults with a RT-PCR-confirmed diagnosis of SARS-CoV-2 infection within 5 days of the original PCR result.

For the *Innova SARS-CoV-2 Antigen Rapid Qualitative Test*, testing was additionally performed for a subset of samples on-site at four COVID-19 testing centres by trained research staff using the “dry swabs” to evaluate “real-life”/diagnostic performance. Dry swabs are those that are not placed into viral transport medium prior to performing the LFD test.

### Community field service evaluation (Phase 4 evaluation)

Wider field service evaluations were performed within a number of UK institutions and settings. These evaluations utilised the *Innova SARS-CoV-2 Antigen Rapid Qualitative Test*. These institutions included a secondary healthcare setting (John Radcliffe Hospital, Oxford), PHE Porton Down, armed forces members (following an outbreak) and in secondary schools (pupils aged 11-18). Evaluations were also undertaken at regional COVID-19 testing centres as part of an NHS Test and Trace service evaluation involving the general public. The John Radcliffe Hospital, Oxford performed an evaluation as part of their asymptomatic staff screening service using the Respiratory Diagnostic Kit Evaluation (‘Red Kite’) study (Research Ethics Committee reference: 19/NW/0730; North West-Greater Manchester South Research Ethics Committee).

### Statistical analyses

Fisher’s exact and chi-squared tests were used to determine non-random associations between categorical variables. Statistical analyses and data visualisation were performed using R version 4.0.3. Sensitivity and specificity and 95% confidence intervals were calculated using the exact Clopper-Pearson method.

## Supporting information

Author Contributions

## Data Availability

Data is available upon request and at the discretion of the corresponding author.

## Acknowledgements

The authors thank the participants and their families affected by COVID-19, NHS doctors and nurses and other medical staff, research scientists and support staff at Public Health England, Porton Down, NHS Test and Trace COVID-19 testing centres staff, the NIHR research network, the University of Birmingham medical school, the University of Oxford medical school, the University of Newcastle medical school, NHS Test and Trace and St John Ambulance.

We would like to thank all members of the UK Lateral flow oversight group in contributing data at a challenging time as listed in the web appendix (appendix page 1)

We would like to acknowledge the Department of Health and Social Care, NIHR, University of Manchester and University of Oxford Biomedical Research Council in funding this study.

Viral stocks were supplied by Dr Julian Druce, Doherty Institute, Queensland University, Australia. The NHS and funders had no role in data collection, analysis or decision to publish.

