## Supplementary Material for "COVID-19: Rapid Antigen detection for SARS-CoV-2 by lateral flow assay: a national systematic evaluation for mass-testing"

**SUPPLEMENTARY MATERIALS**

**Supplementary Methods**

**Pre-clinical evaluation (Phase 2 evaluation)**

Pre-clinical evaluation of candidate LFDs was performed by trained laboratory scientists at Public Health England (PHE) Porton Down. LFDs were evaluated against SARS-CoV-2 spiked positive controls and known negative controls, consisting of saliva collected from healthy adult staff volunteers. Fresh saliva was confirmed as negative when screened for SARS-CoV-2 RNA by RT-PCR.

Unless otherwise stated, all RT-PCR testing was undertaken on the Roche Cobas® 6800 or 8800 system using their proprietary SARS-CoV-2 assay as per manufacturer’s instructions (with off-board lysis using AVL buffer (Qiagen) and 5% Triton-X100 (Sigma Aldrich)). This assay detects ORF-1a/b as a SARS-CoV-2 specific target, and the E-gene as a pan-sarbecovirus target.

Positive and negative samples were processed in Class III and Class I microbiological safety cabinets, respectively. Each sample was tested on each LFD according to the manufacturer’s instructions. For LFDs designed to be used with swab samples, 200uL of saliva was mixed with the LFD extraction buffer directly. To establish preliminary analytical specificity, we tested each LFD against negative samples from 71 healthy volunteers. To establish preliminary analytical limits of detection (LOD), we tested each LFD against saliva spiked with SARS-CoV-2 as follows: Saliva from 15 SARS-CoV-2 PCR-negative individuals (selected from the 71 negative samples described above) was spiked with SARS-CoV-2 virus stock (7.8x10^6^ plaque-forming units [pfu]/mL (VIC/1/2020) and serially diluted in the saliva from each individual yielding 10^5^, 10^4^, 10^3^ and 10^2^ pfu/mL. Fifteen samples per titre were tested. A further dilution series down to 1.22 pfu/ml was performed as part of a positive extended dilution series. Viral copy numbers per titre were quantified by RT-PCR. If applicable, swab comparisons were also performed to ensure the different types of swabs provided were comparable.

Pre-defined and publicly available “prioritisation” criteria to pass on to the next evaluation phase had to be met for LFDs, consisting of (i) a kit failure rate of <10%; (ii) an analytical specificity of ≥97%, and (iii) an analytical LOD of ≥9 of 15 (60%) at 10^2^ pfu/mL, corresponding to a RT-PCR cycle threshold (Ct) of approximately 25; and (iv) lack of cross-reactivity with seasonal coronaviruses to further test analytical specificity. SARS-CoV-2*-*negative saliva samples from five individuals were spiked with seasonal coronavirus strains 229E, NL63 and OC43 at 1:10 dilution and 200µL of each sample analysed on LFDs.

**Retrospective secondary care evaluation (Phase 3a evaluation)**

Evaluation using patient samples retrospectively was started in August 2020 at PHE Porton Down. Samples were obtained from a secondary healthcare setting (Oxford University Hospitals NHS Foundation Trust). Test performance was assessed by testing naso- and oropharyngeal swabs that had been placed in 1ml of viral transport medium (VTM) (Medical Wire). These samples were confirmed SARS-CoV-2-negative or positive by RT-PCR, as follows:

A sample volume of 100µL was tested in Phase 3a LFD evaluations and results were analysed to identify sensitivity and specificity. The sensitivity for viral antigen detection was determined for strata of Ct values and associated viral loads. Kit failures were also recorded. Viral load (in RNA copies/mL) was quantified from Ct values by using a conversion factor obtained using a dilution calibration series of synthetic genomic RNA (*Twist Bioscience)* and a standard curve performed using Altona and Taqpath ORF and S target assays. Viral load conversion to RNA copies/mL was performed using the following equation derived from prior calibration curves, logVL = 11.19-0.304*(delta CT-4.4).

**Community research evaluation (Phase 3b evaluation)**

Witnessed verbal consent by telephone was performed and participants were invited to attend one of fourteen drive-through testing centres in England. Participants were required to return to the COVID-19 testing centre and self-collect paired swab samples, consisting of combined anterior nasal and oropharyngeal swabs (1 stored as a dry swab and 1 swab placed in VTM). Samples were stored at 4°C and analysis was performed at PHE Porton Down within 24 hours. LFDs were evaluated according to the manufacturer’s instructions using “dry swabs” and were also evaluated on VTM samples as per Phase 3a evaluations.

For the *Innova SARS-CoV-2 Antigen Rapid Qualitative Test*, testing was additionally performed for a subset of samples on-site at four COVID-19 testing centres by trained research staff using the “dry swabs” to evaluate “real-life”/diagnostic performance. The paired swab placed in VTM was used to determine viral load through RT-PCR, again at PHE Porton Down. Only individuals who had a SARS-CoV-2 infection confirmed by this paired PCR were included in the analysis. Analyses quantified kit failure rate and sensitivity. Sensitivity was quantified excluding results recorded as void/invalid due to kit failure. The Ct values recorded for the ORF-1 and E-gene generated on the Roche platform and a mean of the two were recorded. Where symptoms were available from participants, further analysis was also performed assessing LFD performance in relation to clinical symptoms.

**Community field service evaluation (Phase 4 evaluation)**

Wider field service evaluations were performed within a number of UK institutions and settings. These evaluations utilised the *Innova SARS-CoV-2 Antigen Rapid Qualitative Test*. These institutions included a secondary healthcare setting (John Radcliffe Hospital, Oxford), PHE Porton Down, armed forces members (following an outbreak) and in secondary schools (pupils aged 11-18). Evaluations were also undertaken at regional COVID-19 testing centres as part of an NHS Test and Trace service evaluation involving the general public. The John Radcliffe Hospital, Oxford performed an evaluation as part of their asymptomatic staff screening service using the Respiratory Diagnostic Kit Evaluation (‘Red Kite’) study (Research Ethics Committee reference: 19/NW/0730; North West-Greater Manchester South Research Ethics Committee). All samples were self-collected, with participants following printed instructions for sampling. The test was then performed and interpreted by an “operator”. The schools, PHE staff and armed forces used an oropharyngeal sample, hospitals used a nasopharyngeal sample, all other field service evaluations utilised anterior nasal and combined oropharyngeal samples.  With the exception of the armed forced outbreak which utilised swabs placed in VTM, dry swabs were used as per manufacturer’s instructions. Analyses were performed to identify kit failure rate, specificity, and viral antigen detection/sensitivity by LFDs. Further analyses were performed to assess LFD performance in relation to user training.

**Lateral flow tests**

All LFD tests were sourced directly from the manufacturer by DHSC. Unless otherwise stated, kits were used as per manufacturer’s instructions, with kits recommending use of a “dry” swab, whereby the swab is not placed into any solutions prior to transfer to the kit buffer. As per the manufacturer’s instruction for use, an invalid kit result, or a kit failure was recorded when an operator did not see a control line on the device within a defined time period. A negative result was recorded where there was evidence of a control line but no test line. A positive result was recorded where there was evidence of a control and test line. Candidate LFDs were assessed under a confidentiality agreement and only the those which cleared all evaluation phases were approached for agreement to publish their identity.

**Supplementary Tables**

*Supplementary Table 1. Results of phase 3a evaluations showing viral detection sensitivity and specificity of LFD tests using biobanked VTM samples from secondary care.*

| Viral load | ORF1 CT | Innova | Abbott | Orient gene | Deepblue | Fortress | SD Bio swab | Surescreen |
| --- | --- | --- | --- | --- | --- | --- | --- | --- |
| >10million | <18 | 3/3 (100.0) | 3/3 (100.0) | 3/3 (100.0) | 3/3 (100.0) | 3/3 (100.0) | 3/3 (100.0) | 3/3 (100.0) |
| 1-10 million | 18-21.5 | 25/25 (100.0) | 28/28 (100.0) | 27/27 (100.0) | 28/28 (100.0) | 28/28 (100.0) | 27/27 (100.0) | 28/28 (100.0) |
| 0.1-1 million | 21.5-25 | 31/33 (93.9) | 33/35 (94.3) | 32/35 (91.4) | 32/35 (91.4) | 31/35 (88.6) | 33/34 (97.1) | 33/34 (97.1) |
| 10,000-100,000 | 25-28 | 23/34 (67.6) | 25/37 (67.6) | 26/37 (70.3) | 23/37 (62.2) | 28/37 (75.7) | 19/36 (52.7) | 16/37 (43.2) |
| 1,000-10,000 | 28-31 | 12/41 (29.3) | 13/42 (31.0) | 5/42 (11.9) | 4/42 (9.5) | 15/42 (35.7) | 7/42 (16.7) | 0/42 (0.0) |
| 100-1,000 | 31-34.5 | 1/37 (2.7) | 1/41 (2.4) | 0/41 (0.0) | 0/41 (0.0) | 2/41 (4.9) | 0/41 (0.0) | 1/40 (2.5) |
| <100 | >34.5 | 0/5 (0.0) | 0/5 (0.0) | 0/5 (0.0) | 0/5 (0.0) | 0/5 (0.0) | 0/5 (0.0) | 0/5 (0.0) |
| Negative samples | na | 0/940 (0.0) | 5/1589 (0.003) | 0/999 (0.0) | 0/1014 (0.0) | 1/1000 (0.001) | 1/996 (0.001) | 1/995  (0.001) |

Supplementary Table 2. Associations between patient features, symptoms, past medical history and LFD result from individuals with COVID-19 positive PCR samples in the Falcon-C19 study.

|  | Total Cohort (n=421) | LFD-Positive (n=312) | LFD-Negative (n=109) | Odds Ratio (95% CI) | p value | p adjusted |
| --- | --- | --- | --- | --- | --- | --- |
| **Patient Features** |  |  |  |  |  |  |
| Age (Median Years) | 33 | 32 | 35 | 1.00 (0.98-1.01) | 0.63 | ns |
| Age=No data | 91 | 63 | 28 |  |  |  |
| Male | 168 | 126 | 42 | 1.08 (0.65-1.77) | 0.78 | ns |
| Female | 169 | 129 | 40 |  |  |  |
| Gender-No data | 84 | 57 | 27 |  |  |  |
| **Current Symptoms** |  |  |  |  |  |  |
| Asymptomatic | 40 | 27 | 13 | 0.66 (0.33-1.38) | 0.25 | ns |
| Abdominal_pain | 19 | 14 | 5 | 0.92 (0.34-2.92) | 0.88 | ns |
| Anosmia | 67 | 43 | 24 | 0.52 (0.29-0.92) | 0.023 | ns |
| Cough | 134 | 106 | 28 | 1.41 (0.86-2.38) | 0.18 | ns |
| Diarrhoea | 15 | 12 | 3 | 0.99 (0.34-3.62) | 0.99 | ns |
| Fever | 80 | 67 | 13 | 1.94 (1.04-3.85) | 0.046 | ns |
| Headache | 138 | 119 | 19 | 2.91 (1.69-5.21) | 0.00019 | 0.0051 |
| Shortness_of_breath_SOB | 57 | 47 | 10 | 1.67 (0.84-3.65) | 0.17 | ns |
| Sore_throat | 82 | 64 | 18 | 1.23 (0.70-2.26) | 0.49 | ns |
| Vomiting | 16 | 12 | 4 | 0.99 (0.34-3.62) | 0.99 | ns |
| Other | 260 | 203 | 57 | 1.70 (1.02-2.82) | 0.04 | ns |
| No data | 59 | 40 | 19 |  |  |  |
| **Past Medical History** |  |  |  |  |  |  |
| Asthma | 40 | 31 | 9 | 1.17 (0.55-2.70) | 0.71 | ns |
| Diabetes | 8 | 6 | 2 | 1.00 (0.23-6.91) | 1 | ns |
| Heart_disease | 9 | 6 | 3 | 0.66 (0.17-3.17) | 0.56 | ns |
| Hypertension | 19 | 13 | 6 | 0.71 (0.27-2.07) | 0.50 | ns |
| Immunosuppression | 2 | 2 | 0 | na | na | na |
| Other_chronic_lung_condition | 5 | 5 | 0 | na | na | na |
| Renal_impairment | 2 | 2 | 0 | na | na | na |
| Steroid_therapy | 4 | 3 | 1 | 1.00 (0.13-20.35) | 1 | ns |
| Not_applicable | 261 | 195 | 66 | 0.95 (0.55-1.59) | 0.84 | ns |
| No data | 57 | 39 | 18 |  |  |  |

Supplementary Table 3: Sensitivity of each assay for detection of viral loads ≥0.1 million (Ct <25)

| LFD | Evaluation | Sensitivity (95% CI) |
| --- | --- | --- |
| Innova | 3a | 93.7 (84.5 – 98.2) |
|  | 3b | 97.6 (91.5 – 99.7) |
| Abbott | 3a | 97.0 (89.5 – 99.6) |
|  | 3b | 91.4 (76.9 – 98.2) |
| Orient Gene | 3a | 95.4 (87.1 – 99.0) |
|  | 3b | 100.0 (90.0 – 100.0) |
| Deepblue | 3a | 95.4 (87.3 – 99.0) |
|  | 3b | 98.5 (91.8 – 100.0) |
| Fortress | 3a | 93.9 (85.2 – 98.3) |
|  | 3b | not complete |
| SD Bioswab | 3a | 93.9 (85.2 – 98.3) |
|  | 3b | not complete |
| Surescreen | 3a | 98.5 (91.7 – 100.0) |
|  | 3b | not complete |

**Supplementary figures**

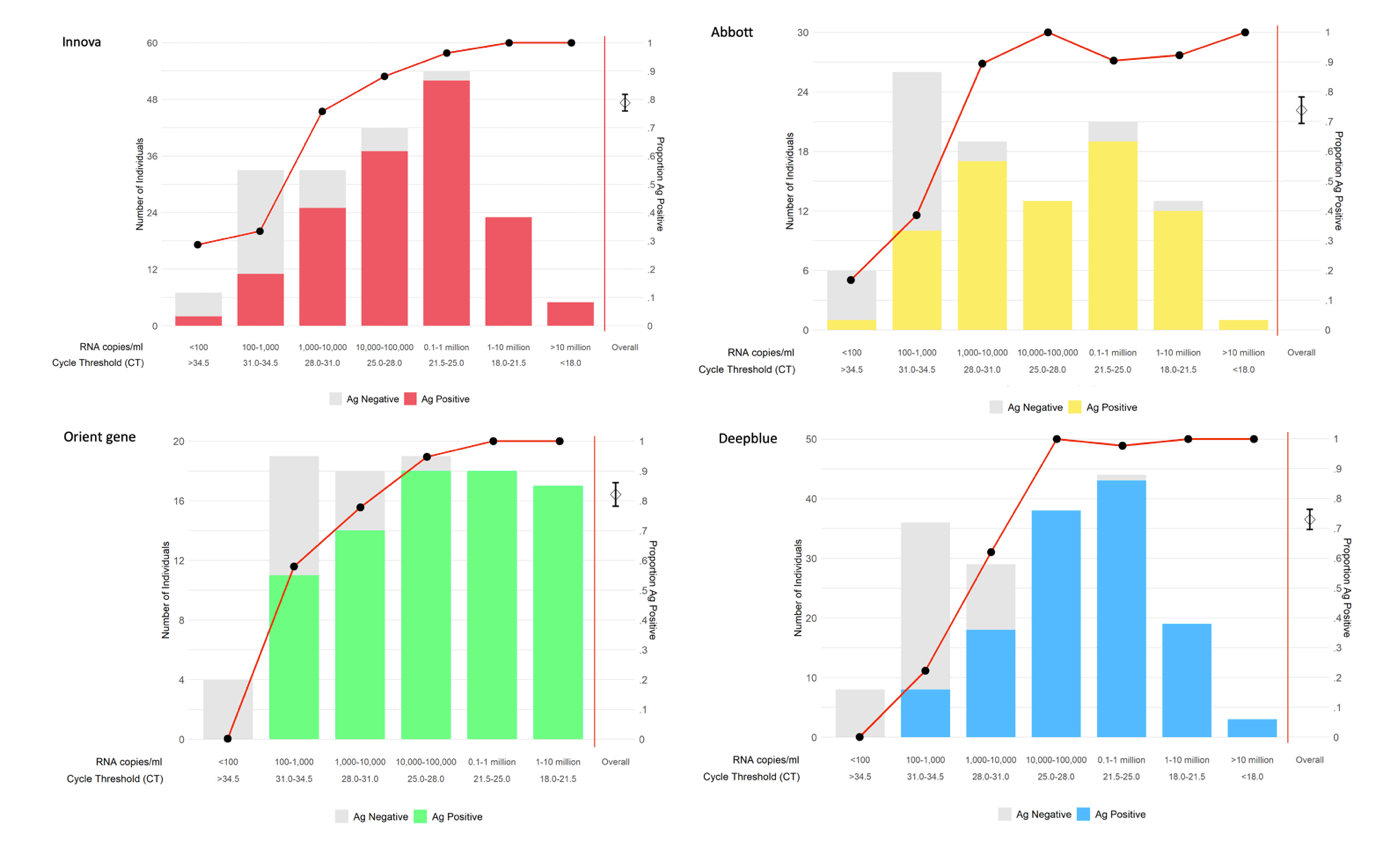

Supplementary figure 1. Sensitivity of LFDs from phase 3b evaluations using dry swabs on Innova, Abbot, Orient gene and Deepblue performed by clinical laboratory personnel.

A	B

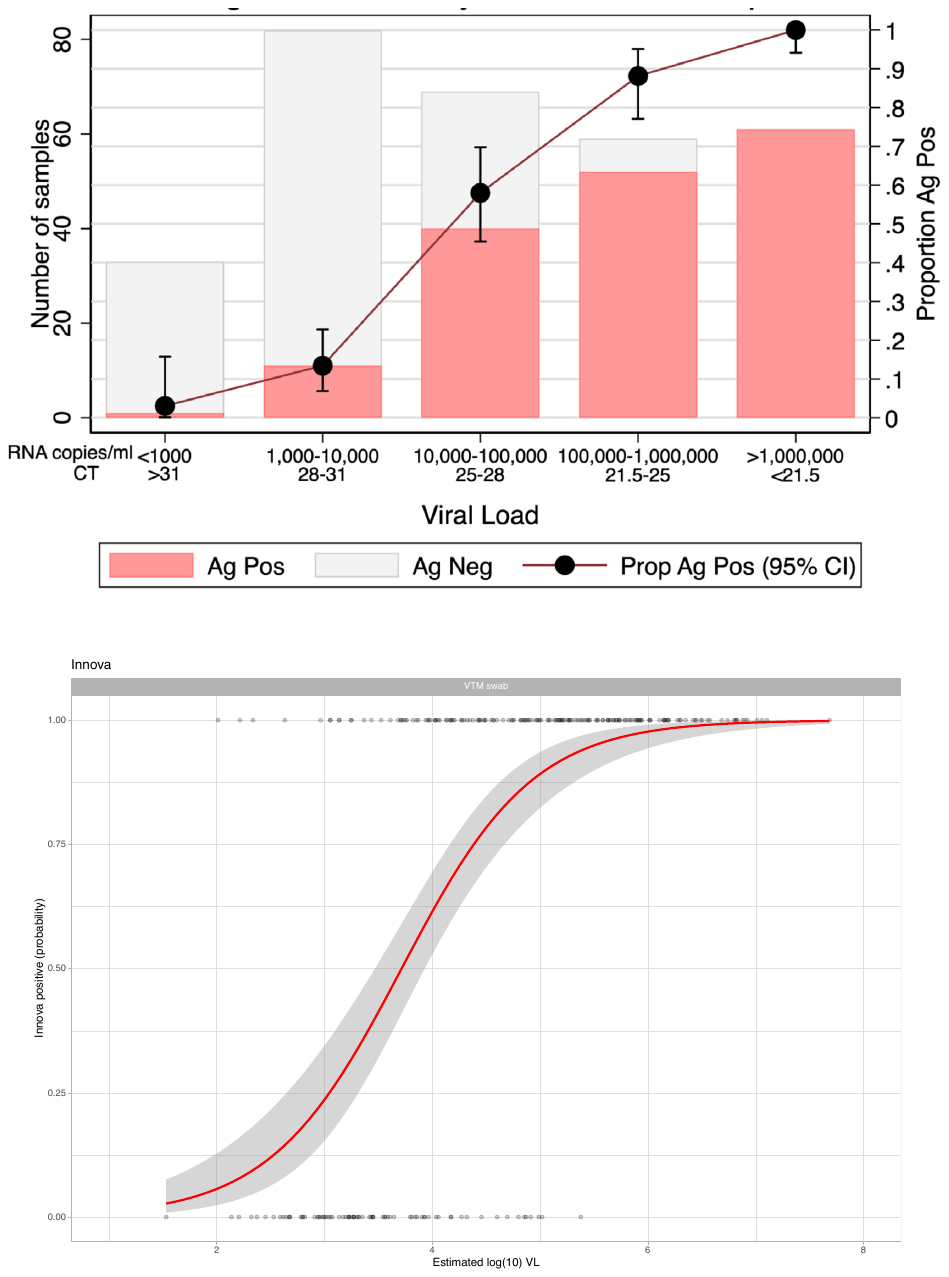

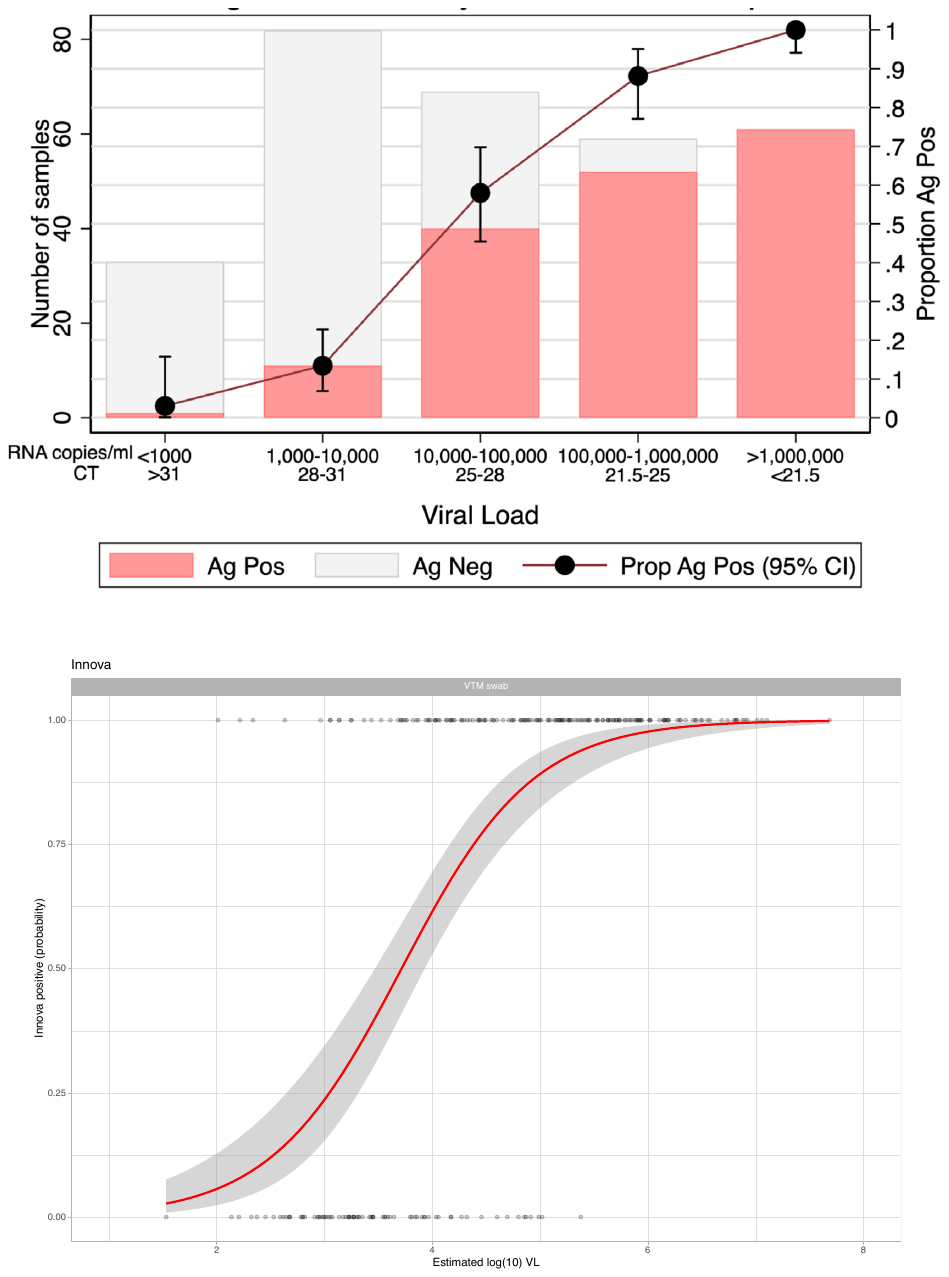

Supplementary Figure 2a. Association between viral antigen detection and viral load (RNA copies/ml and CT) in phase 3b evaluation for swabs taken in VTM when performed by trained laboratory scientists and trained healthcare workers using the Innova test. 2b Modelled probabilities for a positive Innova LFD result at a given log(10) viral load estimate

**Authorship**

This manuscript was prepared by the writing committee and review­ed and approved by all members of the UK COVID-19 lateral flow oversight group.

**Writing Committee (on behalf of the UK COVID-19 lateral flow oversight group)**

Lennard YW Lee, David Eyre, Philippa Matthews, Nicole Stoesser, Abbie Bown, Richard Body, Derrick Crook, Tim Peto

**UK COVID-19 lateral flow oversight group**

John Bell, Derrick Crook, Tim Peto, Lennard YW Lee, Susan Hopkins, Tom Fowler, Jacqui Ramagge, Anna Gronert, Toby Lambert, Alex Sienkiewicz, Hannah Fordham, Mark Driver, Shana Vijayan, Sarah Walker, Emma Stanton, James Bethell

**LFD Evaluation Delivery Group**

Richard Vipond, Neil Mcleod, Abbie Bown, Tim Brooks, Peter Hammond, Cathy Rowe, Bassam Hallis, Lennard YW Lee, Tim Peto, Derrick Crooke, Nicole Stoesser and Phillipa Matthews, Alex Sienciewicz (chair)

**Falcon-C19 project management group**

Daniel Lasserson, Charles Reynard, Lennard YW Lee, Richard Body

**Redkite project management group**

Philippa Matthews, Sheila Lumley, Tim Peto, Nicole Stoesser, Derrick Crook, Sarah Walker.

**Contributions**

**Study conception and design**

John Bell, Richard Body, Abbie Bown, Derrick Crook, Peter Hammond, Lennard Lee, Neil Mcleod, Tim Peto, Alex Sienkiewicz.

**Laboratory tests and evaluation**

*Public Health England Porton Down*

Babak Afrough, Collette Allen, Tracy Benford, Abbie Bown, Tim Brooks, Olivia Carr, Daniel Carter, Matthew Catton, Jim Chadwick, Bea Choi, Simon Clark, Dawn Clarke, Richard Clayton, Ant Crook, Silvia D’Arcangelo, Ruth Elderfield, Mollie French, Joseph Gernon, Bassam Hallis, Peter Hammond, Gabriella Harker, Patrick Lahert, Cristina Leggio, Christopher Logue, Vanessa Lucas, Deborah Fox McKee, Neil McLeod, Ashley Otter, Jodie Owen, Prem Perumal, Cathy Rowe, Jane Shallcross, Angela Sweed, Sian Tiley, Elizabeth Truelove, Richard Vipond, Stephen Wilson, Deborah Wright.

**Data analysis**

Richard Body, Abbie Bown, David Eyre, Peter Hammond, Lennard YW Lee, Philippa Matthews, Neil McLeod, Tim Peto, Cathy Rowe, Alex Sienkiewicz, Thomas Starkey, Nicole Stoesser, Chris Turnbull, Richard Vipond

**Study site set up, logistics and data collection**

*Falcon workstream B study team*

Richard Body, Eloïse Cook, Charles Reynard, Sarah Sampson

*CONDOR steering committee*

Richard Body, Gail Hayward, Philip Turner, Graham Prestwich, Daniel Lasserson, Brian Nicholson, Joy Allen, Pete Buckle, David Price, Mark Wilcox, Kerrie Davies, Val Tate, Julian Braybrook, Anna Halstead, Andrew Lewington, Colette Inkson, Adam Gordon, Paul Dark, Beverley Riley, Rafael Perera-Salazar, Charles Reynard

*Oxford University Hospitals -study management team, Oxford study site and centralised consenting team*

Mark Ainsworth, Susan Bird, Angela Bloss, Wendy Byrne, Penny Carter, Mark Dolman, Joy Edwards, Ranoromanana Evans, David Eyre, Vanessa Fenech, Claire Hall, May Havinden-Williams, Elena Hazell, Susan Johnston, Teresa Lockett, Philippa Matthews, Mike Newbury, Dimitris Papoulidis, Hellen Purnell, Kathryn Saunders, Nicole Stoesser, Ian Wickens, Rangeni Zinyama.

*Manchester team- study management team and Manchester site*

Alison Allanson, Miriam Avery, Carol Beane, Margaret Broughton-Smith, Samatha Chilcott, Richard Clarke, Caroline Coulson, Sally Hammond, Joanne Henry, Heena Mistry, Akhila Muthuswamy, Thanh Pham, Shelha Siddiqui, Anila Sukumaran, Ellie Watson, Louise Woodhead, Bindhu Xavier

*West Midlands team- consenting team*

Pam Devall, Kelly Hollier, David Tyrrell, Juan Dobaldo Pavon, Carly Tibbins, Gurvinder Gill, Jane Mitchell, Jane Wilcocks, Anuradha Krishna, Jake Osbourne Wylde, Jewel Jones Nwanaforo, Sophie Moore.

*West of England CRN*

Geeta Ghadiali, Zoe Lampshire, Shaolin Chidavaenzi, Andrew Harris

*Birmingham study site*

Linda Wagstaff, Victoria Hardy, Lucy Hughes, Wendy Osbourne, Julie Timmins

*Swindon study site*

Suzannah Pegler, Joe Stevens,

*Stoke study site*

Anita Agasu, Mary-Anne Darby, Marion Evans, Emily Eaton, Herika Willis, Steve Hurdowar, Sharon Kempson

*Sheffield study site*

Janet Field, Joanne Jackman, Scott Nicol, Mark Pinkerton, Michelle Platton, Lisa Zeidan, Ella Baldwin

*Peterborough study site*

Lorraine Archer, Marina Bishop, Gloria Calderon, Siobhan Campbell, Rima Colston, Marie Corcoran, Laura Costello, Codie Fahey, Sally-Anne Hurford, Katie Keating-Fedders, Rachel Michel

*The Newcastle-upon-Tyne Hospitals NHS Foundation Trust/Newcastle study site*

Beverley Buck, John Davis, Anika Goel, David Green, Ashley Price, Susan Ridge, Bertie Rowell, Cara Tomas-Smith, Kimerbley Webster, Barbara Wilson, Fiona Yelnoorkar

*Liverpool study site*

James Connolly, Sarah Dyas, Becky Evans, Kerry Gibbons, Gemma Nanson, Hema Thomas, Philip Walker

*Leicester study site*

Natalie Draper, Kate Gilmour Kim Hicklin, Juliet Jones, Karen Pearson, Lucy Sheppard

*Leeds study site*

Carla Bratten, Sally Gordon, Iftikhar Khan, Justin Liu, Sarah Mckee, Julie Miller, Zarina Mirza, Arron Peace, Helen Permain, John Vertannes

*London study site*

Antoinette McNulty

*Coventry study site*

Priya Bagga, Pauline Derbyshire, Kate Ellis, Sue Elwell, Rachel Evans, Narindar Ghuman, Ruth Johns, Sara Read, Jackie Sears, Kate Webberley, Ruth Johns, Katherine Allen, Elaine Butcher.

Gloucestershire Hospitals NHS Foundation Trust

Joanne Waldron, Amanda Selassie, Kayleigh Collins, Jennifer Smith, Kate Trigg-Hogarth, Rachel Sayers, Pauline Brown

*University of Birmingham student trial management team*

Rishab Balaji, Alice Field, Emily Protheroe, Laura Seeney, Amelia Sterry

*University of Birmingham*

Natasha Ashbridge, Iman Aurfan, Ellena Badenoch, Sophie Barraclough, Rosie Boulton, Megan Cathrall, Tanzina Chaudary, Tillie Graham, Tegan Gumsley-Read, Beth Hanney, Arabella Legard, Vindhya Maripuri, Freddie Mellor, Daniel Myers, Sulaksan Panchalingam, Ashley Pegg, Lottie Rawden, James Robinson, Anna Sherkat, Thomas Starkey, Grace Westland.

*University of Oxford student trial management team*

Tinashe Kanyowa, Mika Erik Moeser, Aleksander Stawiarski, Jenny Y Wang

*University of Oxford*

Kevin Chau, Rosaline De Koning, Thomas Foord, Hannah Fuchs, Jasmine Gan, Alexander Grassam-Rowe, Ben Holloway, Sarah Hoosdally, Beinn Khulusi, Kirsten Lee, Sheila F Lumley, Philippa Matthews, Alex Mighiu, Harry Nuttall, Liam Peck, Leon Peto, Hayleah Pickford, Thomas G Ritter, Patrick Robinson, Gillian Rodger, Alexandra Rowlands, Lavanya Sinha, Lloyd Shail, Ella Smith, Nicole Stoesser, Ali Vaughan

*University of Warwick*

Daniel Lasserson

*University of Newcastle*

Hannah Claasen, Harriet Jackson

*Department of Health and Social Care*

Mark Stockbridge, Hannah Fordham, Louise Oliver

*Deloitte**

Dominic Affron, David Chapman, Toi Neibler, Richard Sedgewick, Sami Tatar, Elena Turek.

**Deloitte were not part of the research consortium, but provided support to the consortium in delivering this program.*

*PA consulting*

Louise Oliver

*University Hospitals Birmingham*

Oliver Topping

*Sandwell and West Birmingham Hospitals NHS Trust*

Thomas Knight

*Manchester University NHS Foundation Trust*

Eloïse Cook

*St John Ambulance*

Chris Bridgeman, Dominic Britton, Ben Corley, Graham Ellis, Des Markham, Mary Walters
